# Evaluation of two RT-PCR techniques for SARS-CoV-2 RNA detection in serum for microbiological diagnosis

**DOI:** 10.1101/2020.11.15.20231795

**Authors:** Alexandra Martín Ramírez, Nelly Daniela Zurita Cruz, Ainhoa Gutiérrez-Cobos, Diego Aníbal Rodríguez Serrano, Isidoro González Álvaro, Emilia Roy Vallejo, Sara Gómez de Frutos, Leticia Fontán García-Rodrigo, Laura Cardeñoso Domingo

## Abstract

Presence of SARS-CoV-2 RNA in serum (viraemia) in COVID-19 patients has been related to poor prognosis and death.

The aim of this study was to evaluate the ability of two commercial reverse real-time-PCR (rRT-PCR) kits, cobas® SARS-CoV-2 (Cobas® test) and TaqPath™ COVID-19 CE-IVD RT-PCR Kit (Taqpath™ test), to detect viraemia in COVID-19 patients and their implementation as routine diagnosis in microbiology laboratory.

This retrospective cohort study was conducted with 203 adult patients admitted to Hospital Universitario de La Princesa, (89 Intensive Care Unit and 114 ward) with at least one serum sample collected in the first 48 hours from admission. A total 265 serum samples were included for study.

Evaluation of both rRT-PCR techniques was performed comparing with the gold standard, a Novel Coronavirus (2019-nCoV) Nucleic Acid Diagnostic Kit; considering at least one target as a positive result.

Comparison of Cobas® test and Taqpath™ test with the gold standard method, showed high values of specificity (93.75 and 92.19 respectively) and Positive Predictive Value (92.92 and 99.88 respectively). Nevertheless, sensitivity (53.72 and 73.63 respectively) and Negative Predictive Value (32.53 and 42.99 respectively) were lower; Kappa values were 0.35 for cobas® test and 0.56 for Taqpath™ test.

For both techniques, differences of viraemia detection between the ICU and non-ICU patients were significant (p≤0.001).

Consequently, SARS-CoV-2 viraemia positive results obtained by both rRT-PCR should be considered good tools and may help in handling COVID-19 patients.

Moreover, these methods could be easily integrated in the routine laboratory COVID-19 diagnosis and may open new strategies based on an early COVID-19 treatment.

## Introduction

Coronavirus disease 2019 (COVID-19) is caused by a new type of coronavirus (SARS-CoV-2), that causes a severe acute respiratory syndrome. This novel coronavirus was first described in December 2019 in the city of Wuhan, China (1). At the first of November of 2020, more than a million deaths has been reported worldwide (2).

Patients with COVID-19 may develop mild, moderate or severe symptoms like severe pneumonia, acute respiratory distress syndrome (ARDS) or multiple organ failure (3). To reduce the number of people who may end up with these severe symptoms, it is important to improve the diagnosis and, above all, to find tools that help us predict which patients will have a worse clinical outcome. Since the beginning of the COVID-19 pandemic, the samples that have been used the most for the diagnosis have been those of the respiratory tract. Lately it has been reported that the patient’s serum may be another sample to consider due to the fact that the presence of RNA of SARS-CoV-2 in serum (called viraemia in this study) is related to unfavorable clinical outcomes and multi-organ damage (4, 5). This has been observed also in 2004, with 75% of the patients studied presenting RNA of SARS-CoV in blood samples (6).

The most common tool to detect RNA of SARS-CoV-2 is the real time reverse transcription polymerase chain reaction (rRT-PCR). This technique is highly sensitive, especially if it detects more than two target regions (7). Although most rRT-PCR tests are performed on respiratory tract samples, the genetic material of this novel coronavirus can be detected in other samples also in serum, peripheral blood, feces and other anatomical parts (8). While the faculty of rRT-PCR for respiratory tract samples is well studied, for other kind of samples, as serum, it is not. Therefore, it is important to carry out comparative studies of the different RT-PCRs available to determine the capacity of these techniques to detect SARS-CoV-2 viraemia.

The aim of this study is the evaluation the ability of two commercial rRT-PCR assays (cobas® SARS-CoV-2 Test, Roche Diagnostics, USA and TaqPath™ COVID-19 CE-IVD RT-PCR Kit, Thermo Fisher Scientific, USA), used in daily routine practice of COVID-19 diagnosis, to detect SARS-CoV-2 viraemia; and their implementation in a microbiology laboratory.

## Material and Methods

### Patients and samples

This retrospective cohort study was conducted with 203 adult patients admitted to Hospital Universitario de La Princesa, a tertiary level hospital in Madrid (Spain), between March 1^st^ and April 30^th^, with at least one serum sample collected in the first 48 hours from admission. All patients were symptomatic, with infection by SARS-CoV-2 confirmed by a positive PCR on a nasopharyngeal swab sample. Eighty nine were admitted to the ICU (Intensive Care Unit) with a median age of 65 years (IQR 69-72) and 71.3% male sex; and 114 were admitted to the general ward with a median age of 64 years; (IQR 54.3-72) and 66.4% of them men.

Serum sampling was part of routine clinical practice. Some patients (50) had subsequent serum samples, which were collected during their hospital admission as part of their routine management. A total of 265 serum samples were included for this study. All samples were conserved at −20 °C until they were tested.

### Design of Study

Serum samples were tested with two rRT-PCR: cobas® SARS-COV-2 test (cobas® test), a qualitative assay for detection of SARS-CoV-2 RNA; and TaqPath™ COVID-19 CE-IVD RT-PCR Kit (TaqPath™ test), a multiplex RT-PCR assay for qualitative detection of nucleic acids of SARS-CoV-2. Both are used for routine detection of SARS-CoV-2 from nasopharyngeal swab samples at our hospital.

Results obtained by both techniques were compared with results obtained by Novel Coronavirus (2019-nCoV) Nucleic Acid Diagnostic Kit (Sansure Biotech Inc., China), a multiplex PCR test for qualitative detection of nucleic acids of SARS-CoV-2. This technique was the gold standard method due to it has CE (European Conformity In Vitro Device) and FDA (Food and Drug Administration) authorization for use in blood samples. Sansure Biotech reported a positive agreement of the test of 94.34% (95% CI: 84.34% ∼ 98.82%), and a negative agreement of 98.96 % (95% CI: 96.31% ∼ 99.87 %), in a clinical evaluation performed for the Submission to FDA EUA (Emergency Use Authorization) (9).

### Sample processing

Test assay of all the three rRT-PCR was carried out with 400 µL of serum, treated previously for virus inactivation with lysis buffer.

#### Cobas® test

The assay detects a fragment of the *orf-1ab* region, specific of SARS-COV-2; and a conserved region of *e* gene, a structural enveloped gene, for pan-sarbecovirus detection. Test was performed by cobas® 6800 System (Roche Diagnostics, USA); an automatic platform of nucleic acids extraction and RT-PCR amplification and detection. Serum samples were processed according to manufacturer’s indications, following the same protocol used for SARS-CoV-2 detection in respiratory samples. Results were analyzed and interpreted automatically by the cobas® 6800/8800 Software version 1.02.12.1002.

#### TaqPath™ test and Gold Standard Method test

TaqPath™ test and gold standard method require a previous nucleic acid extraction from sample, which was performed by the automatic eMAG® Nucleic Acid Extraction System (Biomerieux, France). Extraction was carried out according to eMAG® manufacturer’s directions, obtaining purified RNA in 60 µL of elution buffer, which was used to performed both assays.

TaqPath™ test detects three specific SARS-CoV-2 genomic regions: *orf-1ab, s*, and *n* genes and was carried out using 5 µL of purified RNA, according to the manufacturer’s instructions.

Gold standard test detects two specific regions of SARS-CoV-2 genome: *orf-1ab* and *n* genes. The nucleic acid amplification was performed according to the kit manufacturer’s indications using 10 µL of purified RNA.

Both rRT-PCR tests were performed by QuantStudio™ 5 Real Time PCR System (Applied Biosystems, USA). Amplification curves were analyzed with QuantStudio™ Design and Analysis software version 2.4.3 (Applied Biosystems, USA). Interpretation of results were done by a clinical microbiologist, means of the amplification curves analysis, considering for a positive target detection: a) a Cycle threshold (Ct) cutoff value of 40; b) curves with typical S-shape or without plateau.

### Analysis of results

Detection of at least one target was considered as a positive result.

Evaluation of viraemia in patients was carried on considering only the results obtained from their first serum sample.

Results obtained from all the 265 samples collected were analyzed for the techniques assessment.

Statistical analysis was performed using SPSS 25.0 (IBM Corp., USA). Continuous and categorical variables were presented as median (interquartile range, IQR) and n (%), respectively. To analyze differences between detection of viraemia by each technique in patients, according if they have been admitted at ICU or not, a χ2 test was performed. The Ct of any detected target was recorded and it was calculated the median, interquartile range (IQR) and the 95^th^ percentile for each one. Sensitivity, specificity, positive and negative predictive values and Cohen’s Kappa coefficient were calculated for cobas® test and TaqPath™ test, in comparison with the gold standard method.

## Results

### Viraemia detection in patients

The cobas® test detected viraemia in 50,2% of patients recruited, showing a 65,2% and 38,6 % of positive viraemia detection for ICU and non-ICU admitted patients, respectively. On the other hand, TaqPath™ test detected viraemia in 62,6% of of all patients recruited, with a viraemia detection in 75,3 % of patients admitted in ICU, and 52,6% in those patients not admitted at ICU had a positive result. For both techniques, differences of viraemia detection between the ICU and non-ICU patients were significant (p≤0.001). Results are shown in Table 1.

**Table 1:** Viraemia detection in patients admitted at ICU or in ward by both evaluated techniques.

|  |  | Ward (%) | ICU (%) | Total | p |
| --- | --- | --- | --- | --- | --- |
| **Cobas® SARS-CoV-2** | Negative | 70 (61.4) | 31 (34.8) | 101 (49.8) | 0.0002 |
|  | Positive | 44 (38.6) | 58 (65.2) | 102 (50.2) |  |
|  | **Total** | **114** | **89** | **203** |  |
| **TaqPath™ COVID-19** | Negative | 54 (47.4) | 22 (24.7) | 76 (37.4%) | 0.001 |
|  | Positive | 60 (52.6) | 67 (75.3) | 127 (62.6) |  |
|  | **Total** | **114** | **89** | **203** |  |

p value calculated by $\chi^2$ test

Furthermore, the gold standard assay was able to detect the presence of RNA SARS-Cov-2 in 80,8% of patients, with the following distribution: 85.4% and 77.2% for ICU or Non-ICU patients, respectively. No significant differences between both groups were found (p=0.14).

### Assessment of serum samples

A total of 265 serum samples were analyzed by both rRT-PCR methods: cobas® test and TaqPath™ test.

Comparison between cobas® test and the gold standard showed a 65.66% of concordance, with a kappa coefficient of 0.35. On other hand, comparison of TaqPath™ test showed a concordance of 78.11%. A kappa value of 0.52 was obtained. Summarized results of sensitivity, specificity, positive and negative predictive values of both techniques are shown in Table 2.

**Table 2:**
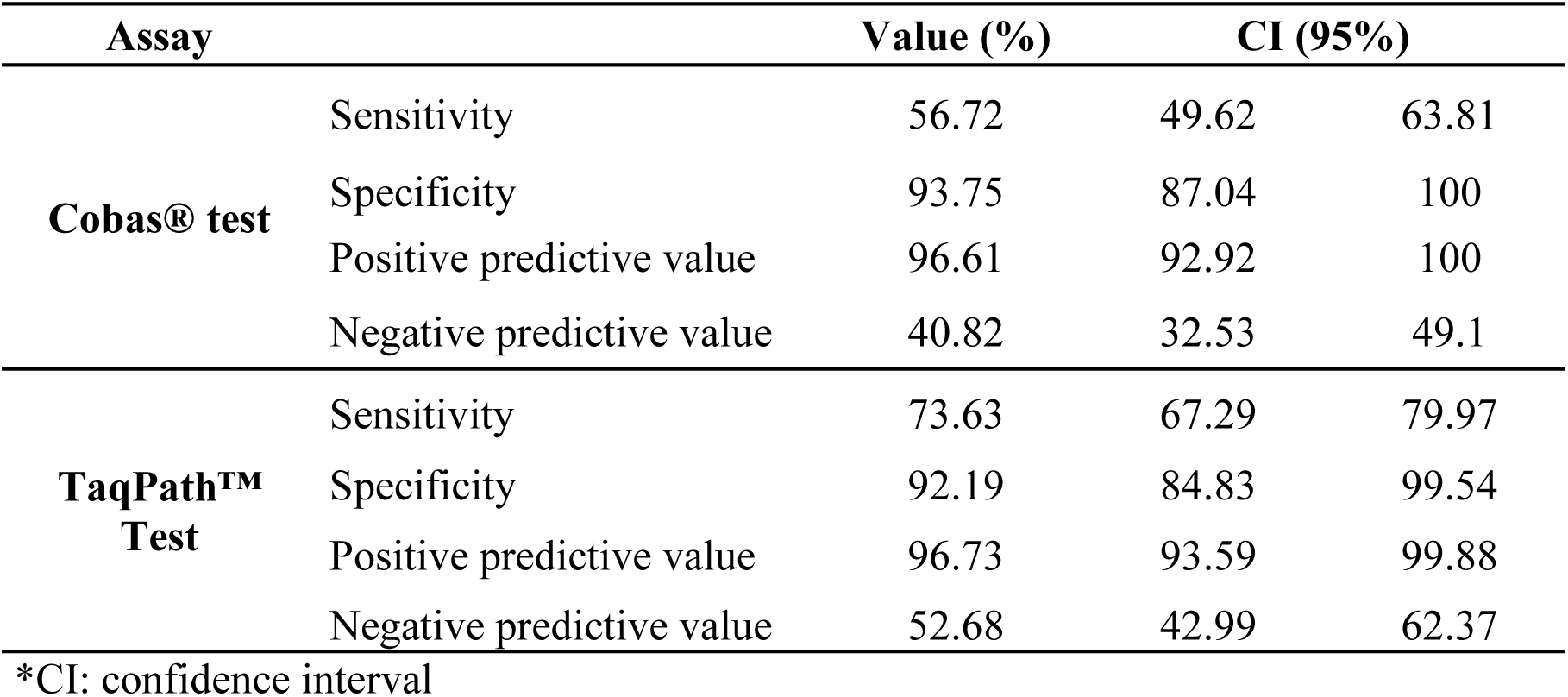
Sensitivity, Specificity, positive and negative predictive values and their confidence intervals (95%) of cobas® test and TaqPath™ test compared to the gold standard method.

Median Ct value of cobas® test targets detected were 36.2 (IQR 34.2-37.1) for *e* gene, and 32.7 (IQR 32.1-34.1) for *orf1ab* gene. The 95^th^ percentiles were 38.72 and 35.21 for *e* and *orf1ab* genes, respectively. On the other hand, median Ct values for each target detected by TaqPath™ test were: 31.91 (IQR 29.92-33.27); 32.46 (30.65-34.31) and 31.81 (30.01-32.92) for *s, n* and *orf1ab* genes respectively, and the 95^th^ percentile of Ct was 36.91, 37.17 and 37.04 for *s, n* and *orf1ab* genes respectively. Ct values of TaqPath™ test are shown in Table 3.

**Table 3:**
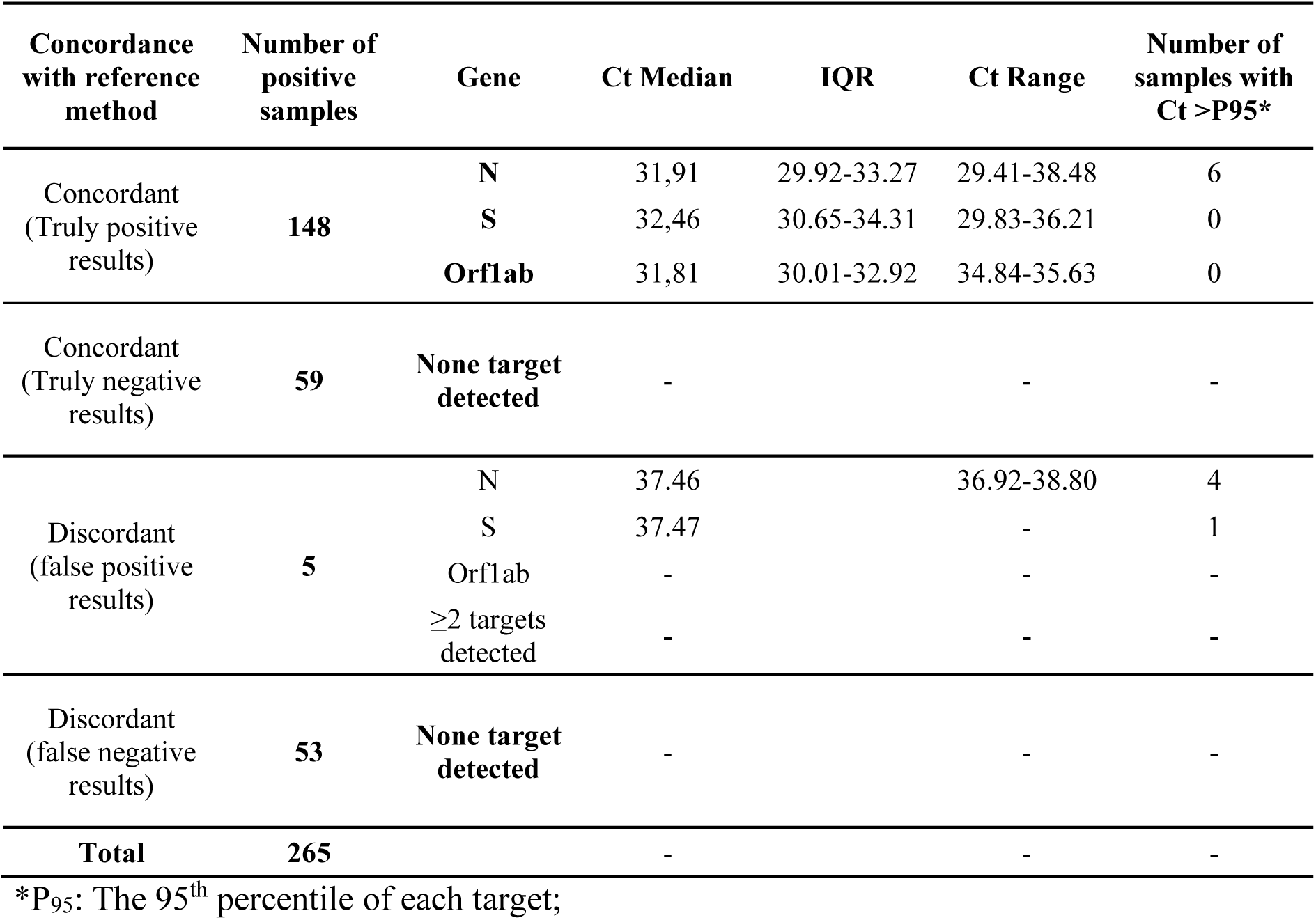
Number of samples detected with TaqPath™ test in comparison with the gold standard method.

| Concordance with reference method | Number of positive samples | Gene | Ct Median | IQR | Ct Range | Number of samples with Ct >P95* |
| --- | --- | --- | --- | --- | --- | --- |
| Concordant (Truly positive results) | 148 | N | 31,91 | 29.92-33.27 | 29.41-38.48 | 6 |
|  |  | S | 32,46 | 30.65-34.31 | 29.83-36.21 | 0 |
|  |  | Orf1ab | 31,81 | 30.01-32.92 | 34.84-35.63 | 0 |
| Concordant (Truly negative results) | 59 | None target detected | - |  | - | - |
| Discordant (false positive results) | 5 | N | 37.46 |  | 36.92-38.80 | 4 |
|  |  | S | 37.47 |  | - | 1 |
|  |  | Orf1ab | - |  | - | - |
|  |  | ≥2 targets detected | - |  | - | - |
| Discordant (false negative results) | 53 | None target detected | - |  | - | - |
| **Total** | **265** |  | - |  | - | - |

\*P<sub>95</sub>: The 95<sup>th</sup> percentile of each target;

Cobas® test obtained 4 false positive results with only detection of *e* gene and with Ct values from 37.57 to 38 while by TaqPath™ test 5 false positives were detected by detection of one target gene (*n or s* genes) only, and the Ct ranged from 36.92 to 38.8.

False negative results were obtained by cobas® test for 87 samples, 30 of them with amplification of the two targets of the gold standard method; the rest of false negative samples (57/87) showed amplification of just one target gene. On the other hand, 53 false negatives were detected by TaqPath™ test; most of them (42/53) were positive by detection of only one target gene of the gold standard method, meanwhile the rest

(11/53), were positive by detection of both, n and orf1ab genes. There was agreement in 45 (85%) of the false negatives obtained by TaqPath™ test with cobas® test.

## Discussion

To our knowledge, this is the first study which assesses the performance and accuracy of these techniques in serum samples for SARS-CoV-2. The goal of this study is to assess two commercial rRT-PCR assays for SARS-CoV-2 RNA detection in serum samples, to provide clinicians a useful tool in the management of the COVID-19. Although these commercial assays have not been validated for their use in serum or plasma samples, the extraction methods performed in this study are commonly carried out in serum or plasma samples for other viruses (10–12).

The preferred method for diagnosis of SARS-CoV-2 is rRT-PCR of upper respiratory tract samples, via nasopharyngeal and oropharyngeal swabs or, in some cases, lower respiratory tract samples (13). However, SARS-CoV-2 have been detected in multiple samples, as saliva, stool, plasma and serum (8, 14). Detection of SARS-CoV-2 RNA in serum samples have been related to progression to a critical disease and death (4, 15) and with the cytokine storm (3). However, most of the studies perform in-house or commercial methods for SARS-CoV-2 detection are not validated in serum or plasma samples (4, 15).

In our study the viraemia’s percentage showed significant differences (p≤0,001) in ICU patients respect patients admitted in the ward with the two rRT-PCR assays; supporting previous studies which indicate that presence of SARS-CoV-2 RNA in serum samples is more common in critical patients and could be considerated a prognostic indicator (15–17).

Comparison between the assessed rRT-PCR kits and the gold standard method showed high specificity and positive predictive values, over 90% in both techniques. These high values point out that a positive result of RNA detection of SARS-CoV-2 in serum samples of patients previously COVID-19 diagnosed are reliable, and should be taken into account for clinicians attending to the patient.

Cobas® test showed four false positive results, all of them with the only amplification of *e* gene. According to manufacturer’s indications, detection of the *e* gene without amplification of *orf-1ab* region could be due to several factors, mutations in the amplification region of *orf-1ab*, low viral loads, presence of other sarbecovirus or other factors. These results should be considered as presumptive positive results and be retested (18). Moreover, 52 true positive samples had amplification of the *e* gene, so it was decided to consider the amplification of only this target as positive for several reasons. First, because the serum samples collected in this study belonged to previously diagnosed COVID-19 patients. Secondly, according to the WHO (19), SARS-CoV has only been reported just four times since the end of the epidemic on July 2003 (three times associated to laboratory accidents and another one in Southern China). So the probability of the samples with *e* gene amplification without any other targets are due to other sarbecovirus is very low, and other reports considered them as positive too (10).

There were five false positive results with TaqPath™ test, all of them with just one target detected and with high Ct values (from 36.92 to 38.80). Even more, these Ct values were over the 95^th^ percentile, which could suggest the probability to obtain these false positive results is low. Therefore, values over 37 are unlikely, and should not be considered, which is in agreement with the last manufacturer’s indications (20). However, with this consideration, six concordant results considered as true positive would turn into false negative results, decreasing sensitivity slightly to 72.68% but increasing specificity to 98.33% (supplementary table 1).

On the other hand, the negative predictive values obtained with both cobas® test and TaqPath™ test were low (40.82% and 52.68%, respectively), due to the high number of false negative results obtained (87 with cobas® Test and 53 with TaqPath™ test). Moreover, it is remarkable that there was a high coincidence of the false negative results between both assessed techniques, 85% TaqPath™ test being negative with cobas® Test.

It is difficult to elucidate the reason of these false negative results, as the three employed rRT-PCR kits are commercial and the design is not available for the customers, presenting differences in the number of targets, and possibly in the sequence or the size of the amplicons, which cause differences in sensitivity of the techniques. Even more, the cobas® test is performed in the automatic close cobas® 6800 system, making more difficult to elucidate the nature of discrepancies. A possible reason is that these samples had low viral loads, decreasing the sensitivity of the assessed techniques. Also, the RNA eluted volume added to the PCR reaction could have influenced because it was the double with the gold standard method compared with the TaqPath™ test.

Moving to onboarding to the clinical practice, both techniques can be easily implemented in the diagnosis of COVID-19 in the microbiology laboratory and they can be performed along with the rest of respiratory samples for the diagnosis of SARS-CoV-2. TaqPath™ test is more suitable performing a small number of tests at a time meanwhile the cobas® test should be more useful when it is necessary to analyze high amounts of samples

The present study has some limitations. First, the intrinsic analytical variability of PCR can have influenced in the results obtained.

Another limitation is the type of sample used in this study. Although serum samples are accepted in the analysis of viraemias, the common samples used in most microbiology laboratories are plasma samples.

In conclusion, this study shows that cobas® SARS-CoV-2 Test and TaqPath™ COVID-19 CE-IVD RT-PCR Kit could be useful in SARS-CoV-2 viraemia detection in serum of diagnosed COVID-19 patients. For a better test performance, considering the detection of at least one target obtained with any technique, and in particular with a Ct value under 37 with the TaqPath™ assay. A viraemia positive result may help in handling COVID-19 patients, due to the relationship between viraemia and bad prognosis and mortality recently reported (4, 15). In addition, these techniques are easy to incorporate in the Microbiology laboratory routine for SARS-CoV-2 diagnosis.

Finally it could open new treatment strategies based on early COVID-19 treatment (15).

## Data Availability

All data referred in our manuscript are available to see for every one who wants to do it.

## Compliance with Ethical Standards

### Funding

The authors did not receive any fundings to do the study.

### Ethical approval

All participants enrolled voluntarily, and written informed consent was required to use data for analysis. The stydy was approved by the Hospital Universitario La Princesa independent ethics research committee (reference number 4267, acta CEIm 21/20).

---

**Supplementary table 1.** Comparison values calculated considering the Ct> P_95_ as a negative result.

|  | TaqPath™ COVID-19 Kit | cobas® SARS-Cov-2 Test |
| --- | --- | --- |
| Sensitivity | 72.68% | 55.84% |
| Specificity | 98.33% | 93.75% |
| Positive predictive value | 99.3% | 96.49% |
| Negative predictive value | 52.68% | 40.82% |
| Kappa coefficient | 0.55 | 0.35 |

